# Temporal inequalities in the global COVID-19 vaccine rollout: a cross-national observational study of delivery, health-system capacity, and time to coverage

**DOI:** 10.64898/2026.08.29.26361724

**Authors:** Hsuan-Wei Lee, Yi-Hsuan Huang, Thomas McAndrew

## Abstract

**Introduction:** By the end of 2023, many low-income countries had not reached 50% COVID-19 vaccine coverage, while most high-income countries had exceeded 80%. It remains unclear whether receiving vaccine deliveries translated into faster population coverage. We examined cross-national inequalities in the timing of the vaccine rollout and whether deliveries through the COVID-19 Vaccines Global Access (COVAX) facility were associated with subsequent national uptake.

**Methods:** We conducted an observational study of 218 countries and territories using country-level data up to December 2023. We used generalized additive mixed models to identify country-level correlates of coverage at an early and a later stage of the pandemic, survival analysis to compare the time to 50% coverage between COVAX Advance Market Commitment (AMC) and non-AMC countries, and an event study to estimate the association between the timing of the first COVAX delivery and subsequent monthly coverage in AMC countries.

**Results:** AMC-supported countries reached 50% coverage substantially more slowly than non-AMC countries. The hazard of reaching the threshold was 0.17 times that of non-AMC countries at month 1 (95% CI 0.07 to 0.41) and 0.53 times at month 18 (95% CI 0.33 to 0.85). One year after rollout began, 65.9% of AMC countries (95% CI 56.7 to 76.6) had not reached 50% coverage, compared with 21.1% of non-AMC countries (95% CI 15.1 to 29.5). The timing of COVAX deliveries was not significantly associated with subsequent national uptake in any post-delivery month. In the early stage of rollout, higher maternal mortality was associated with lower coverage, while a larger urban population was associated with higher coverage. By the end of the observation period, larger household size was associated with lower coverage, while higher health expenditure and a larger urban population were associated with higher coverage.

**Conclusion:** Receiving COVAX deliveries was not, on its own, associated with faster coverage. Coverage differences were more consistently associated with country-level structural and health-system characteristics, while we found no significant association with the timing of the first COVAX delivery. Achieving vaccine equality likely requires strengthening the capacity of health systems to convert deliveries into administered doses, and preparedness efforts should invest in last-mile delivery capacity ahead of future emergencies.

**Key messages:** *What is already known on this topic:* Low-income countries lagged far behind high-income countries in COVID-19 vaccine coverage, and although COVAX was created to narrow this gap, few studies examined whether receiving deliveries translated into faster population uptake.

*What this study adds:* Across 218 countries and territories, AMC-supported countries reached 50% coverage substantially more slowly than non-AMC countries, and the timing of COVAX deliveries showed no significant association with subsequent national uptake. Coverage was more consistently associated with country-level structural factors, namely maternal mortality and urban population in the early stage and household size, health expenditure and urban population by the end of the period.

*How this study might affect research, practice or policy:* Vaccine delivery alone may be insufficient to accelerate coverage. Preparedness efforts should invest in the health-system capacity needed to convert deliveries into administered doses, and should measure success by how quickly populations are protected rather than by doses shipped.

## 1. Introduction

The COVID-19 pandemic exposed deep differences in how quickly countries were able to protect their populations through vaccination. By the end of 2023, more than 13 billion doses had been administered worldwide, yet coverage remained highly uneven: most high-income countries passed 80 percent coverage within about a year, while many low- and middle-income countries had not reached half of their populations ^1,2^. These differences had real consequences during periods of high transmission, when vaccination was among the few tools available to reduce severe disease and death ^3^.

Throughout this paper we describe these differences as inequalities rather than inequities: we compare the timing and level of coverage across countries without adjusting for underlying need. A full assessment of inequity would require accounting for differences in need, most importantly population age structure, the dominant driver of COVID-19 mortality risk^4^; that is beyond the present scope and we return to it as future work.

To reduce supply-side inequality, the COVID-19 Vaccines Global Access (COVAX) facility was created to pool procurement and channel donated doses to lower-income countries. A central part of this architecture was the Advance Market Commitment (AMC), a financing mechanism that supported 92 low-income and middle-income economies that could not fully self-finance vaccine purchases ^5–7^. Countries eligible for this support are referred to here as AMC countries. COVAX delivered billions of doses to these countries, yet coverage in AMC-supported countries consistently lagged behind wealthier nations^8–10^.

Much of the existing evidence documents the size of the coverage gap at a single point in time or within a particular region ^2,11^. Fewer studies ask a more operational question, namely whether the arrival of deliveries was actually followed by faster population uptake. This distinction matters because supplying doses and administering them depend on different things. Delivery depends on procurement and logistics, while administration depends on cold chain capacity, health workforce availability, and public willingness to be vaccinated ^12–14^. Studies that examine only a single time point, or that pool countries without following them relative to when their own deliveries arrived, cannot cleanly separate these two stages. An approach that follows each country around the timing of its first delivery is better suited to this question.

In this study we examine cross-national inequalities in the COVID-19 vaccine roll-out across 218 countries and territories using three complementary approaches. We first identify the country-level structural characteristics associated with coverage at an early and a later stage of the pandemic. We then compare how quickly AMC and non-AMC countries reached 50 percent coverage. Finally, we estimate whether the timing of the first COVAX delivery was associated with subsequent national uptake in AMC countries. Together these analyses ask not only how large the inequalities were, but whether supplying vaccines was, on its own, associated with narrowing them.

## 2. Methods

This study applies a three-part analytical framework: Generalized Additive Mixed Models (GAMMs) for country-level correlates of coverage at an early and a later stage, survival analysis for the time to reach 50% coverage in AMC versus non-AMC countries, and an event study for the association between the timing of the first COVAX delivery and subsequent uptake, with descriptive stratification by World Health Organization (WHO) region and Global Health Security (GHS) Index category.

### 2.1 Generalized Additive Mixed Models

To examine which country-level characteristics were associated with COVID-19 vaccine uptake, we employed Generalized Additive Mixed Models (GAMMs), which accommodate flexible covariate relationships and, through random effects, the hierarchical structure of the data. In our specification the smooth term enters as a regional random effect rather than as nonlinear smooths of continuous covariates, so the substantive predictors are modelled parametrically^15,16^.

The models were fitted with the bam() function in the R package mgcv; the full model equations are given in the Supplementary File.

Two models were constructed based on data from different pandemic phases: one using data from July 2021, reflecting the early stage of vaccine rollout, and the other using data from 2023, representing the later stage of the pandemic. This temporal division allowed us to assess whether the factors influencing vaccine uptake differed across pandemic phases. The dependent variable was the adjusted partial-vaccination share, defined as a proportion bounded between 0 and 1. The model included structural and policy covariates spanning household composition and size, population and settlement, educational attainment and public spending on education, urbanisation, health-system capacity, and a monthly government-response index.

These covariates were systematically selected to represent five underlying dimensions of national development status: socio-demographic structure, healthcare capacity, human capital, spatial geography, and global supply. These five dimensions, the variables assigned to each, and the associated collinearity diagnostics are set out in the Supplementary File (Supplemental Table S6). To ensure model stability against multicollinearity, Variance Inflation Factors (VIF) were assessed for all baseline structural covariates across both stages. Confirming the absence of severe collinearity, all baseline main-effect VIF values remained safely below the conservative threshold of 5 (Stage 1 maximum VIF = 3.82 for household size category; Stage 2 main-effect maximum VIF = 4.26 for household size category).

To mitigate multicollinearity among the three variables reflecting AMC participation, post-AMC status and lagged cumulative COVAX deliveries, we combined them into a single principal component (PC1), which captured 72.1% of their variance and loaded on all three with the same sign; full loadings are reported in the deposited output. PC1 was sign-oriented so that higher values indicate greater AMC participation and COVAX-related support. Because AMC eligibility also reflects differences between lower- and higher-income countries, PC1 should be read as a composite gradient of AMC participation and its correlates, not as an isolated measure of COVAX support. PC1 was entered as a single predictor, and regional heterogeneity was modelled with WHO-region random intercepts. The models used a quasibinomial family with a logit link, estimated by restricted maximum likelihood with automatic smoothing selection. In the Results, each predictor is summarised by its average marginal effect on the response scale, while the p-values and significance markers refer to the corresponding model terms.

### 2.2 Survival Analysis

Building on the GAMM estimates, we used survival analysis to examine temporal disparities in rollout: whereas GAMMs capture coverage at fixed points in time, survival analysis models the time until a country reaches 50% uptake. The event of interest was the date on which a country’s uptake first exceeded 50%, as recorded in the Our World in Data (OWID) database. We also compared overall per-capita vaccine accessibility and pandemic impact between AMC and non-AMC countries using Wilcoxon rank-sum tests.

The survival time for each country was measured in days, starting from the date the country first recorded any COVID-19 vaccine uptake. The one-year landmark was defined as 365.25 days, the mean calendar year length. Countries were divided into two groups based on whether they were classified as Gavi COVAX Advance Market Commitment (AMC) countries, following the World Health Organization (WHO) classification and the May 2021 COVAX report (Supplemental Table S2). The 88 AMC countries analysed here are those of the 92 AMC-eligible economies that are represented in the country-level series used in this study; the four remaining eligible economies do not appear in that series and are therefore not included in any of the analyses.

Countries that had not achieved the 50% threshold by December 2023 were treated as right-censored, allowing direct comparison of rollout timing between AMC and non-AMC countries. The resulting estimates describe inequalities in timing; any relationship with geopolitical or institutional differences between the groups is beyond what this comparison can establish.

The proportional hazards assumption was assessed using Schoenfeld residuals and was modestly violated (global *p* = 0.011). We therefore also fitted a flexible parametric (Royston-Parmar) survival model with a time-varying coefficient for AMC status as a complementary analysis. The test result and residual plot (Supplemental Table S1 and Supplemental Figure S1), and the full specification, diagnostics and estimates of the flexible model, are reported in the Supplementary File.

### 2.3 Event Study Analysis

Complementing the GAMM and survival analyses, we applied an event study design to countries supported by the Gavi COVAX Advance Market Commitment (AMC) to estimate the temporal association between COVAX vaccine deliveries and subsequent vaccine uptake, allowing a granular assessment of how deliveries coincided with shifts in population-level coverage.

In this study, the event of interest is defined as the month of the first recorded COVAX vaccine delivery in each AMC country. The outcome is national partial-vaccination coverage, taken as the last value recorded in each calendar month and expressed in percentage points on a 0 to 100 scale. For participating countries that received multiple sequential COVAX shipments over time, the temporal benchmark (*t* = 0) was strictly anchored to the arrival date of the first delivery. Our design is a within-group staggered-adoption event study rather than a difference-in-differences setup with an explicit control group. Identification comes from differences in delivery timing across AMC countries, so the interpretation of these temporal associations relies on the parallel pre-treatment trends assumption. The pre-delivery coefficients were not statistically distinguishable from zero (Table 4). Although this does not establish parallel trends, it provides no evidence of differential pre-delivery trajectories within the available event window.

The core model estimates the dynamic associations of vaccine delivery using a series of event time indicators (leads and lags), controlling for time-invariant country characteristics and global time trends.

Countries were stratified by WHO World Region and Global Health Security (GHS) Index ^17^ to capture cross-regional and preparedness-level heterogeneity. The WHO regional distribution comprised AFRO (*N* = 32), AMRO (*N* = 10), EMRO (*N* = 9), EURO (*N* = 5), SEARO (*N* = 7), and WPRO (*N* = 12). Since the GHS Index is continuous, we grouped countries into three fixed GHS bands: low (0 *≤* GHS *≤* 20, *N* = 4), moderate (20.1 *≤* GHS *≤* 40, *N* = 63), and high (40.1 *≤* GHS *≤* 60, *N* = 7) preparedness levels (with 1 country excluded due to missing GHS data). Among AMC countries only these three bands, spanning scores from 0 to 60, were populated. This allowed us to explore heterogeneity in vaccine uptake trajectories across institutional and regional contexts following COVAX delivery.

The empirical analysis focused primarily on AMC countries due to their relatively slower uptake patterns. By the end of 2023, 51% of AMC countries had reached the 50% vaccine uptake threshold, in contrast to 83% among non-AMC countries (Table 1). This disparity, supported by survival analysis findings, motivated further exploration of how international aid and delivery logistics were associated with temporal vaccine accessibility. The analytic sample for each component of the study is summarised in Figure 1.

**Figure 1:**
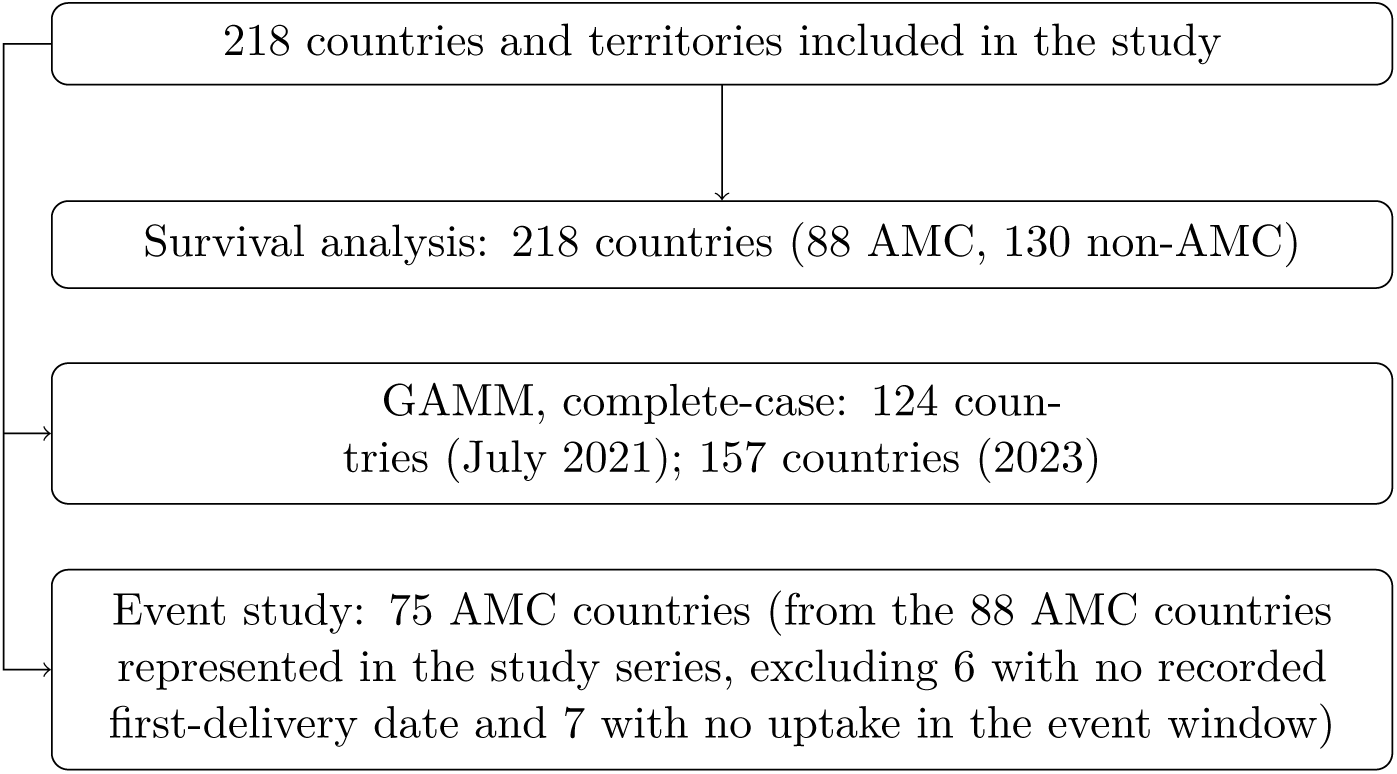
Analytic sample for each component of the study. The three analyses use different samples drawn from the 218 countries; the counts are not nested subsets.

**Table 1:** COVID-19 vaccine uptake (50% threshold) among AMC and non-AMC countries as of 2023.

| Group | Achievement of 50% Threshold | N | Percentage |
| --- | --- | --- | --- |
| AMC | Yes | 45 | 51% |
| AMC | No | 43 | 49% |
| Non-AMC | Yes | 108 | 83% |
| Non-AMC | No | 22 | 17% |

The event study framework enabled estimation of uptake trajectories before and after COVAX delivery, offering insights into the temporal dynamics and timing of multilateral vaccine interventions in low- and middle-income settings.

### 2.4 Data Sources and Collection

This study utilizes a variety of globally recognized datasets to investigate the factors influencing COVID-19 vaccine uptake at the country level. The primary source of demographic and socio-economic data is the GLOPOP-S dataset ^18^, a large-scale synthetic global population database that includes information from over one billion households as of 2015. Household-level attributes such as age, gender, education, household type, settlement type (urban/rural), and household size were aggregated to country-level indicators. Specifically, the mean values were calculated for continuous variables (e.g., rural population share), while the mode was used for categorical attributes (e.g., household type, education level, household size) to capture the most representative characteristics for each country.

Additional country-level data were compiled from authoritative global sources, including the World Health Organization (WHO), Our World in Data (OWID), the Oxford COVID-19 Government Response Tracker (OxCGRT), the World Bank, the Global Health Security Index (GHS Index), and the United Nations Children’s Fund (UNICEF). Vaccine uptake trajectories, pandemic impact metrics (cases and deaths per million) and AMC-related supply metrics were sourced from the OWID and WHO databases. Total vaccine doses delivered, used only for the Wilcoxon accessibility tests, came from the UNICEF COVID-19 Market Dashboard. Government response indicators, including the stringency index and a vaccine-prioritisation variable used only in a sensitivity analysis (Supplemental Table S7), came from OxCGRT. Health expenditure, maternal mortality and public education spending came from the World Bank. Definitions of the model variables and their transformations are provided in the Supplementary File, while the underlying variable names and coding are documented in the deposited replication files. All external data were accessed between December 2024 and March 2025. All variables underwent consistency checks and cross-validation across datasets.

To account for the evolving nature of the pandemic, two time-specific datasets were constructed. The first dataset corresponds to July 2021, representing the early stage of global vaccine rollout, covering 124 countries. The second dataset, representing the later stage in 2023, includes 157 countries. Countries with missing values in key predictors were excluded so that models were estimated on complete-case data, leaving only countries with complete records at each time point.

For the event study analysis, we restrict the sample to 75 AMC countries, drawn from the 88 AMC countries represented in the study series by excluding 13 for temporal alignment: six had no documented initial COVAX delivery date, and seven lacked overlapping vaccination records within the relative event-time window of [*−*5, 12] months.

#### Patient and public involvement

Patients and the public were not involved in the design, conduct, or reporting of this study, which used only aggregated country-level secondary data.

## 3. Results

### 3.1 Generalized Additive Mixed Models

The early-stage model (July 2021) explained 64.2% of deviance (adjusted *R*^2^ = 0.600) across 124 countries.

The results (Table 2) indicate that a one-unit increase in log-transformed urban population was associated with higher predicted vaccine uptake (coefficient *p* = 0.003; AME = 0.171, 95% CI: 0.064 to 0.279), suggesting that countries with larger urban populations tended to achieve higher levels of partial vaccination during the early rollout period. In contrast, a one-unit increase in maternal mortality ratio was associated with lower predicted vaccine uptake (coefficient *p* = 0.007; AME = −0.0008, 95% CI: −0.0014 to −0.0002), highlighting the importance of underlying health system capacity in facilitating vaccine rollout.

**Table 2:** Summary of Generalized Additive Mixed Model (GAMM) Results for Partial COVID-19 Vaccine Uptake (July 2021)

| Variable | Estimate | Std. Error | t value | p-value | AME<br>[95% CI] |
| --- | --- | --- | --- | --- | --- |
| (Intercept) | -3.521 | 2.988 | -1.178 | 0.241 | — |
| Dominant household size category (log, +1) | -1.193 | 1.167 | -1.023 | 0.309 | -0.211<br>[-0.615, 0.193] |
| Dominant educational attainment level (log, +1) | 0.539 | 0.329 | 1.636 | 0.105 | 0.095<br>[-0.018, 0.209] |
| Total population (log, +1) | -0.070 | 0.050 | -1.400 | 0.164 | -0.012<br>[-0.030, 0.005] |
| Population density (log, sqrt, +1) | 0.153 | 0.120 | 1.276 | 0.205 | 0.027<br>[-0.014, 0.069] |
| Government response stringency (log, +1) | 0.176 | 0.252 | 0.699 | 0.486 | 0.031<br>[-0.056, 0.119] |
| Maternal mortality ratio | -0.005 | 0.002 | -2.773 | **0.007\*\*** | **-0.0008\*\***<br>[-0.0014, -0.0002] |
| Public spending on education (% of GDP, sqrt) | 0.113 | 0.229 | 0.493 | 0.623 | 0.020<br>[-0.059, 0.099] |
| Urban population (log, +1) | 0.969 | 0.316 | 3.063 | **0.003\*\*** | **0.171\*\***<br>[0.064, 0.279] |
| COVAX participation and supply (PC1) | 0.007 | 0.069 | 0.104 | 0.917 | 0.001<br>[-0.023, 0.025] |
| *Smooth term (Random effect)* |  |  |  |  |  |
| WHO regional |  | edf = 2.628 | F = 0.907 | 0.138 | — |

Adjusted $R^2 = 0.600$ , Deviance explained = 64.2%, Scale est. = 0.12668, $n = 124$ .
Note: The Estimate, Std. Error, t value and p-value columns refer to the parametric coefficients on the logit scale. Average marginal effects (AME) give the average change in the predicted probability of partial vaccine uptake (0–1 scale) for a one-unit increase in the covariate as it enters the model, that is in its log-transformed or square-root-transformed value. AME standard errors and 95% confidence intervals [CI] are obtained by the delta method, and the interval rather than a separate p-value summarises AME precision. Significance markers: . $p < 0.10$ , \* $p < 0.05$ , \*\* $p < 0.01$ , \*\*\* $p < 0.001$ .

No statistically significant associations were observed for household size, dominant educational attainment, total population, population density, government response stringency, public spending on education, or the composite measure of AMC participation and COVAX support (PC1).

The smooth effect of WHO region was not statistically significant (*p* = 0.138), indicating no residual geographic variation after accounting for the country-level characteristics in the model (Table 2). Residual diagnostics are reported in the Supplementary File (Supplemental Figure S5).

Building upon the findings from July 2021, the analysis was extended to the end of the COVID-19 data collection period, employing a similar GAMM framework to capture how the correlates of vaccine uptake had changed. The dependent variable remained the adjusted partial vaccination rate. This model included interaction terms between population density and both health expenditure and stringency index, as well as an interaction between public education spending and rural population share, to better reflect the complex socio-demographic and policy environment.

The results (Table 3) indicate that a one-unit increase in log-transformed household size was associated with a substantial decrease in predicted vaccine uptake (coefficient *p* = 0.002; AME = −0.532, 95% CI: −0.852 to −0.212). This finding suggests that countries with larger household structures tended to experience lower vaccine uptake, highlighting potential household-level barriers in accessing vaccination. Similarly, a one-unit increase in log-transformed health expenditure was associated with higher predicted vaccine uptake (coefficient *p* = 0.001; AME = 0.122, 95% CI: 0.039 to 0.204), underscoring the importance of health system capacity in facilitating vaccine access.

**Table 3:** Summary of Generalized Additive Mixed Model (GAMM) Results for Partial COVID-19 Vaccine Uptake (End of Data Collection)

| Variable | Estimate | Std. Error | t value | p-value | AME<br>[95% CI] |
| --- | --- | --- | --- | --- | --- |
| (Intercept) | -2.373 | 2.619 | -0.906 | 0.366 | — |
| Household type:<br>couple with children<br>and (non-)relatives | 0.003 | 0.614 | 0.005 | 0.996 | 0.021<br>[-0.102, 0.144] |
| Household type: one<br>parent with children<br>and (non-)relatives | -1.699 | 1.103 | -1.540 | 0.126 | -0.305 .<br>[-0.666, 0.056] |
| Dominant household<br>size category (log, +1) | -2.587 | 0.806 | -3.211 | **0.002\*\*** | **-0.532\*\***<br>[-0.852, -0.212] |

Table 3 continued from previous page
| Variable | Estimate | Std. Error | t value | p-value | AME<br>[95% CI] |
| --- | --- | --- | --- | --- | --- |
| Dominant educational attainment level (log, +1) | 0.237 | 0.299 | 0.793 | 0.429 | 0.049<br>[-0.072, 0.169] |
| Urban population (log, +1) | 0.496 | 0.212 | 2.334 | **0.021\*** | **0.102\***<br>[0.017, 0.187] |
| Population density (log, sqrt, +1) | 0.955 | 0.775 | 1.233 | 0.220 | 0.022<br>[-0.027, 0.071] |
| Health expenditure (log, +1) | 2.308 | 0.702 | 3.286 | **0.001\*\*** | **0.122\*\***<br>[0.039, 0.204] |
| Government response stringency (log, +1) | -0.488 | 0.343 | -1.424 | 0.157 | 0.031 .<br>[-0.004, 0.066] |
| Public spending on education (% of GDP, sqrt) | 0.618 | 0.395 | 1.566 | 0.120 | 0.042<br>[-0.031, 0.114] |
| Rural population share (log, +1) | 2.555 | 2.385 | 1.072 | 0.286 | -0.089<br>[-0.398, 0.219] |
| Population density × Health expenditure | -0.810 | 0.309 | -2.622 | **0.010\*\*** | — |
| Population density × Government stringency | 0.302 | 0.163 | 1.859 | 0.065 . | — |
| Public education spending × Rural population | -1.482 | 1.081 | -1.371 | 0.173 | — |
| *Smooth term (Random effect)* |  |  |  |  |  |
| Household type × WHO region | | edf = 7.254 | $F = 5.370$ | **<0.001\*\*\*** | — |

Table 3 continued from previous page
| Variable | Estimate | Std. Error | t value | p-value | AME<br>[95% CI] |
| --- | --- | --- | --- | --- | --- |

**Table 4:** Event-study estimates of monthly COVID-19 vaccine coverage around the first COVAX delivery.

| Event Time | Estimate | Std. Error | t-value | p-value |
| --- | --- | --- | --- | --- |
| -5 | 3.986 | 6.117 | 0.652 | 0.517 |
| -4 | 3.268 | 4.554 | 0.718 | 0.475 |
| -3 | 2.398 | 2.873 | 0.835 | 0.407 |
| -2 | 1.720 | 1.393 | 1.235 | 0.221 |
| 0 | -1.237 | 1.210 | -1.022 | 0.310 |
| 1 | -2.854 | 1.814 | -1.573 | 0.120 |
| 2 | -1.671 | 2.353 | -0.710 | 0.480 |
| 3 | -2.827 | 2.743 | -1.031 | 0.306 |
| 4 | -2.120 | 2.963 | -0.715 | 0.480 |
| 5 | -2.071 | 3.008 | -0.689 | 0.493 |
| 6 | -1.529 | 2.971 | -0.515 | 0.608 |
| 7 | -3.263 | 3.395 | -0.961 | 0.340 |
| 8 | -1.431 | 3.007 | -0.476 | 0.636 |
| 9 | -0.433 | 2.714 | -0.160 | 0.874 |
| 10 | -1.676 | 2.473 | -0.678 | 0.500 |
| 11 | -0.485 | 1.917 | -0.253 | 0.800 |

*Note:* Event time $t = -1$ serves as the omitted reference category. Month $t = 12$ is excluded due to exact collinearity with year-month fixed effects. Standard errors are clustered at the country level. Estimates are in percentage points of coverage.
*Sample Power Distribution:* The number of unique countries contributing to each event-time coefficient is 75 for months $-5$ to $6$ , and 74 for months $7$ to $11$ .

The interaction between population density and health expenditure was significantly negative (*p* = 0.010), indicating that the positive association between health expenditure and vaccine uptake was attenuated in countries with higher population density. The interaction between population density and government response stringency did not reach conventional statistical significance (*p* = 0.065), although the point estimate was positive, which would be consistent with a stronger association between government response and uptake in more densely populated countries.

The smooth term for household type by WHO region was highly significant (*p <* 0.001), indicating substantial geographic heterogeneity in vaccination patterns across household structures. The adjusted *R*^2^ of 0.571 and deviance explained of 58.9% suggest a moderate model fit. Residual diagnostics for this model are reported in the Supplementary File (Supplemental Figure S6). Figure 2 sets the coefficient estimates from the two stages side by side.

**Figure 2:**
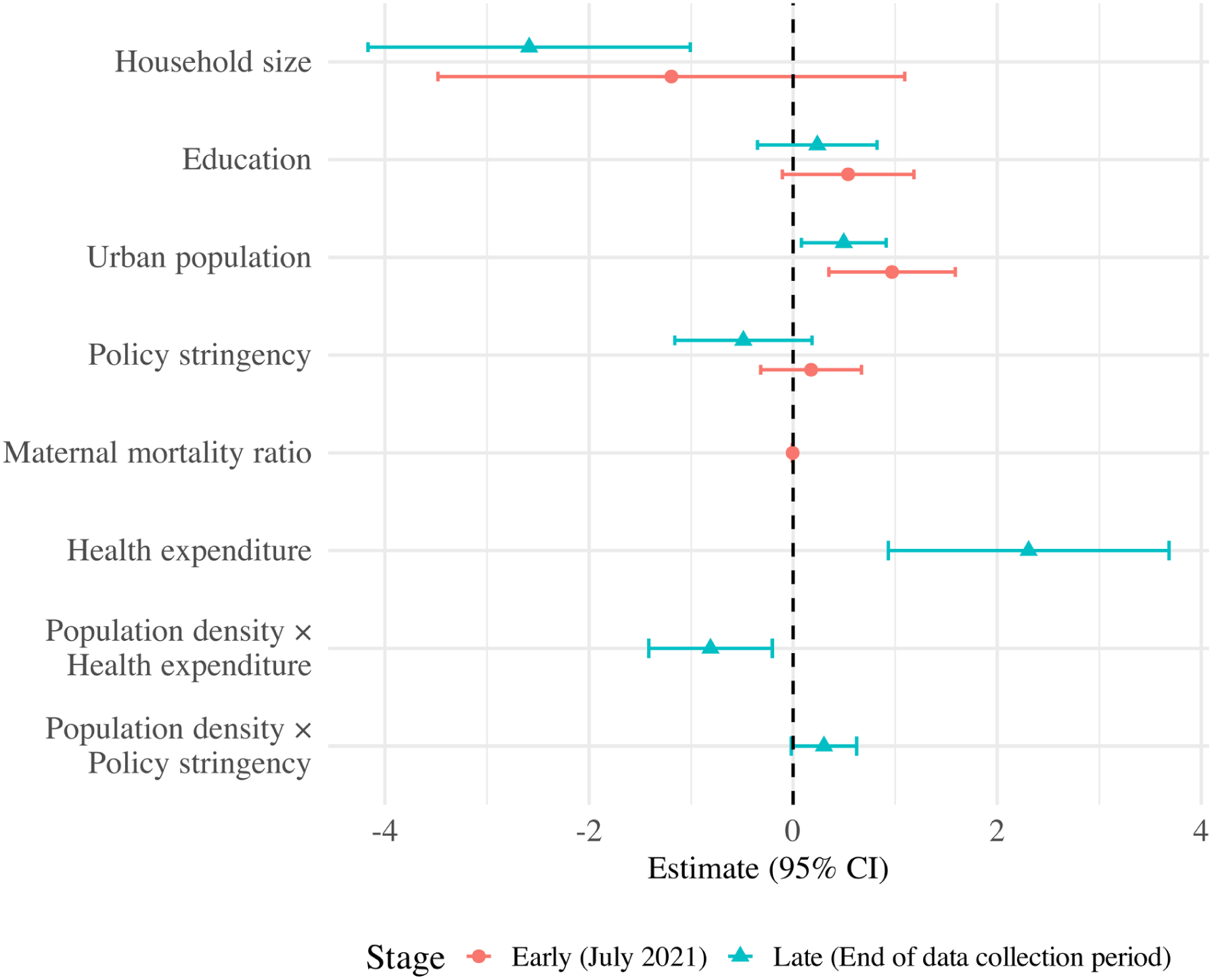
Forest plot of Generalized Additive Mixed Model (GAMM) estimates across two stages. Points show the parametric coefficients on the logit scale and horizontal bars show the corresponding 95% confidence intervals; the early stage refers to July 2021 (124 countries) and the late stage to the end of the data collection period (157 countries). Variable names are abbreviated, and each covariate enters the model as specified in Tables 2 and 3, including any logarithmic or square-root transformation. For readability the figure displays a subset of the model terms; the complete set of parametric coefficients, standard errors, average marginal effects and smooth terms is reported in those tables. The estimates are associational and are not interpreted as causal.

### 3.2 Event Study Analysis

Across the full observation period, non-AMC countries achieved consistently higher COVID-19 vaccine uptake than AMC-supported countries, and this pattern held within every WHO region, with the widest dispersion among AMC-supported countries (Supplemental Figure S2).

Figure 3 presents the estimated dynamic associations between the first COVAX delivery and national COVID-19 vaccine uptake, based on an event study specification. The reference period is defined as one month prior to the first COVAX delivery (event time = *−*1), allowing for a direct comparison of pre- and post-delivery trajectories. Country fixed effects and year–month fixed effects are incorporated to account for time-invariant country-specific characteristics and common temporal shocks, thereby adjusting for potential confounding and examining the temporal trajectory. The results reveal no statistically significant increase in uptake during the post-delivery period. All point estimates after delivery remain close to zero with confidence intervals crossing zero, indicating that coverage did not depart from the trajectory implied by the country and calendar-time fixed effects. This is compatible with factors beyond delivery timing, such as local distribution capacity, shaping subsequent uptake, although our design does not measure those directly.

**Figure 3:**
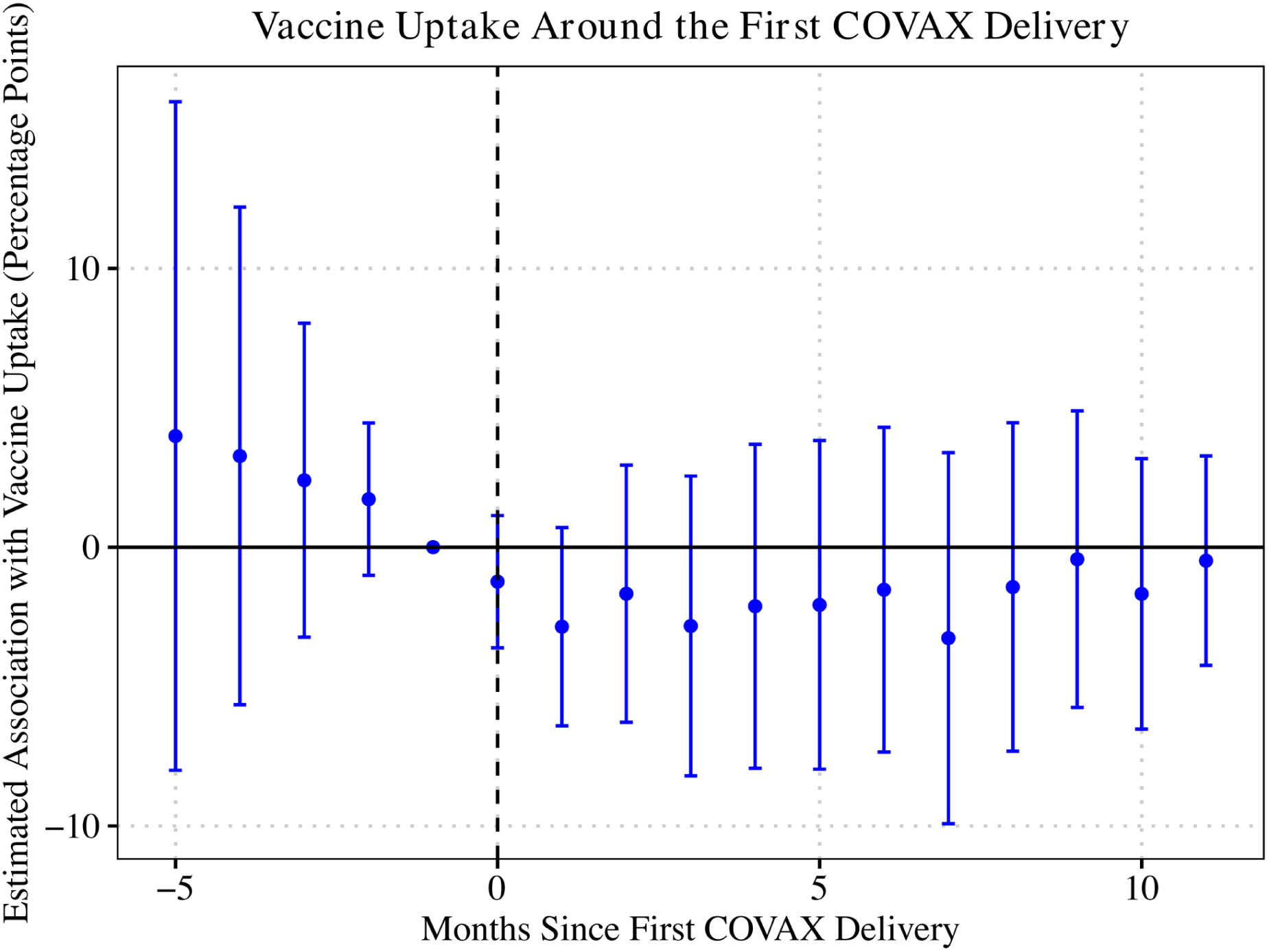
Estimated association between the timing of the first COVAX delivery and subsequent vaccine uptake. The reference period is set to one month before the first COVAX delivery (event time = *−*1). Country and year-month fixed effects are included to control for time-invariant country characteristics and common temporal shocks. All estimated associations are reported in percentage points of coverage, and the horizontal axis is event time in months relative to the first COVAX delivery.

Table 4 reports the event-study estimates for COVID-19 vaccination coverage around the first COVAX delivery across 75 participating countries. Regarding model diagnostics, the within-*R*^2^ of 0.00552 indicates that event-time indicators account for a very small fraction of the within-unit variance in monthly vaccine uptake, while fixed effects absorb the vast majority of overall variation. The results show no significant increase during the entire period after vaccine delivery, a pattern that held across alternative daily and monthly specifications (Supplemental Table S8). The estimated coefficients remain very small and are not statistically different from zero. For example, the estimate is *−*1.237 percentage points in the month of delivery (*t* = *−*1.022, *p* = 0.310) and *−*1.529 percentage points by the sixth month (*t* = *−*0.515, *p* = 0.608).

Together, the low within-*R*^2^ and null estimates suggest that receiving COVAX shipments did not automatically speed up vaccination once global time trends and country fixed effects were accounted for, leaving most within-country variation unexplained and compatible with a role for other factors such as distribution capacity. We next examined whether trajectories differed by health-security capacity and region.

Following the first COVAX delivery, uptake trajectories were ordered by health-security capacity, with the lowest-capacity group declining most steeply and reaching the lowest values after month seven. Because these subgroups are small and the trajectories are not centred on the reference month, we describe this ordering without attaching statistical inference to it (Supplemental Figure S3).

Uptake trajectories also varied across WHO regions, with regions such as SEARO and WPRO tending to remain near baseline while others declined after delivery.

### 3.3 Survival Analysis

A total of 218 countries and territories were included in this study, encompassing both AMC-supported and non-AMC-supported nations. Of these, 153 countries achieved a 50% vaccine uptake rate within the observation period, whereas 65 did not.

Figure 4 presents Kaplan–Meier curves for the time to reach 50% vaccine uptake in AMC-supported and non-AMC countries, with time measured in days. The curves show non-AMC countries reaching the threshold more rapidly than AMC-supported countries. At the start of follow-up, 130 non-AMC and 88 AMC-supported countries were at risk of reaching the threshold; the non-AMC group left the risk set faster, consistent with their earlier attainment of 50% coverage.

**Figure 4:**
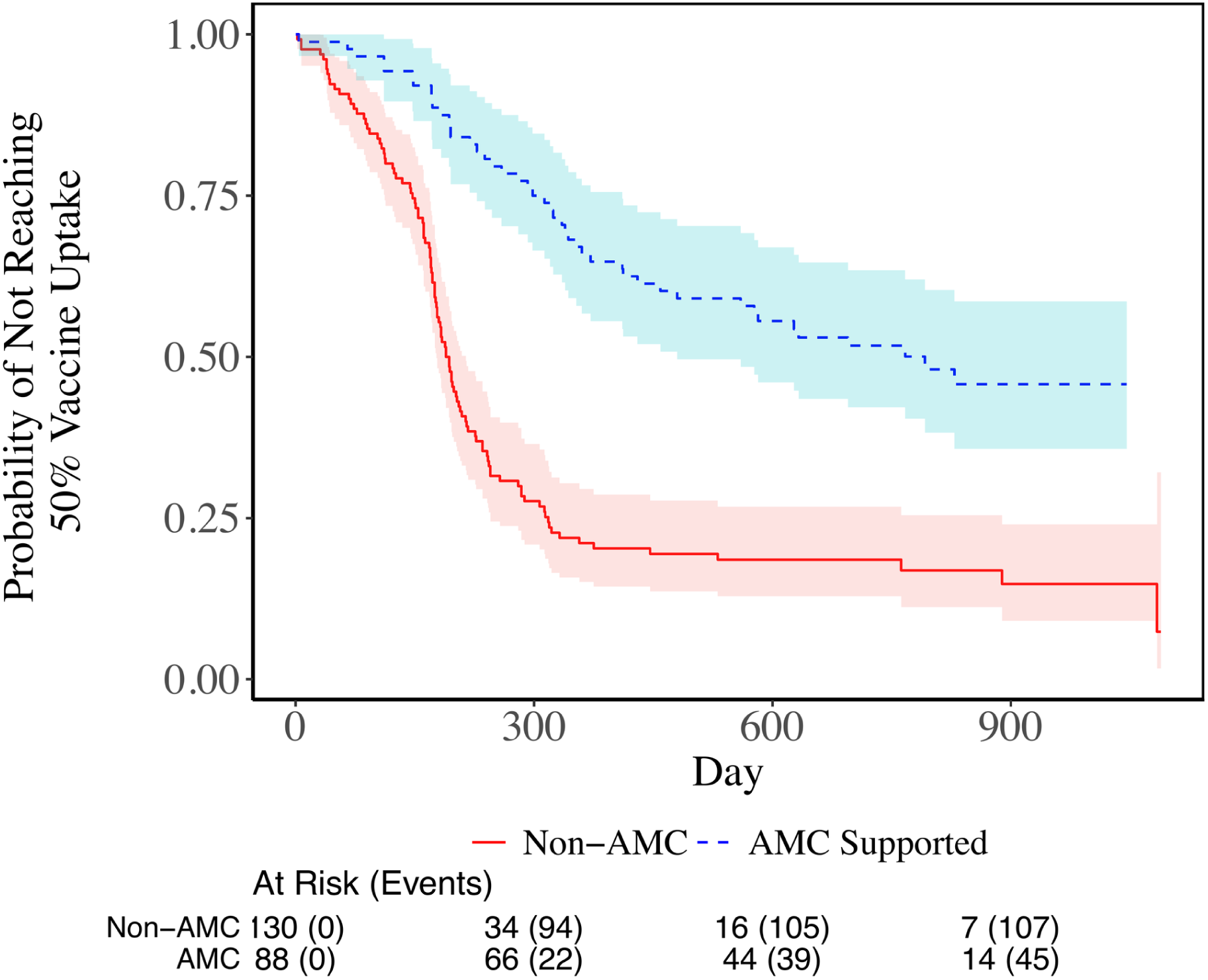
Kaplan–Meier survival curves comparing the time to reach 50% COVID-19 vaccine uptake between AMC-supported and non-AMC countries. The y-axis represents the proportion of countries that have not yet reached the 50% threshold, with time measured in days. The red solid line represents non-AMC supported countries, while the blue dashed line represents AMC supported countries. The risk table below the figure indicates the number of countries remaining at risk at each time point, providing additional context for the observed differences between groups.

The log-rank test, reported in Supplemental Table S3, confirms a statistically significant difference between the two groups (*χ*^2^ = 42.9*, p* = 6*e−* 11), indicating that non-AMC countries were more likely to achieve 50% uptake in a shorter time. At the time of one year (365.25 days) in Supplemental Table S4, the estimated proportion of countries that had not yet reached 50% COVID-19 vaccine uptake was 21.1% for non-AMC countries (95% CI: 15.1%–29.5%) and 65.9% for AMC-supported countries (95% CI: 56.7%–76.6%) as shown in Supplemental Table S4, indicating that non-AMC countries reached the 50% threshold more rapidly.

A flexible parametric survival model with a time-varying coefficient for AMC status told the same story from a different angle. AMC-supported countries were slower to reach the threshold early in the rollout, and this relative difference narrowed over the follow-up period. The full model estimates and the time-varying hazard ratio are reported in the Supplementary File (Supplemental Table S5 and Supplemental Figure S4).

Wilcoxon rank-sum tests showed that AMC-supported countries had significantly lower per-capita vaccine acquisition than non-AMC countries. The gap was already present in 2021 (median 78.5 versus 171.0 doses per 100 population; *W* = 1734, *p* = 5.03 *×* 10^−12^) and persisted to the end of the period (130.0 versus 230.0; *W* = 2136, *p* = 6.17 *×* 10*^−^*^9^). Confirmed cases and deaths per million also differed significantly between the groups, although we do not interpret the direction of those comparisons because recorded cases and deaths depend heavily on national testing and death-registration capacity.

## 4. Discussion

This study set out to measure inequalities in the global COVID-19 vaccine rollout and to ask whether the arrival of COVAX deliveries was associated with faster population coverage. Three findings stand out. First, AMC-supported countries reached 50 percent coverage much more slowly than non-AMC countries, and a large share had still not reached that threshold a year after beginning their rollout. Second, once we followed AMC countries around the timing of their own first delivery, the arrival of COVAX supply showed no significant association with subsequent monthly coverage. Third, the country-level characteristics associated with coverage were structural rather than the timing of supply itself: a larger urban population and lower maternal mortality in the early stage, and a larger urban population, smaller household size and higher health expenditure in the later stage.

Read together, these results point away from a simple supply story: deliveries did not translate automatically into faster coverage, and differences between countries tracked features of their health systems and social structure. This is consistent with the view that supplying doses is necessary but not sufficient, and that delivery and administration capacity may have been an important constraint for many countries. We interpret these associations cautiously, as the design is observational and countries receiving deliveries earlier or in larger volumes may differ systematically from others in ways our models do not capture.

The flexible parametric survival analysis adds a temporal dimension: the gap in the hazard of reaching 50 percent coverage was widest early in the rollout and narrowed over the following months, a pattern compatible with differences in delivery readiness rather than supply alone. Our data do not identify why the gap narrowed.

Several associations confirm existing literature rather than adding novelty: the link between larger household size or higher maternal mortality and lower coverage is consistent with decades of research on the social and health-system determinants of immunisation ^14,19–21^. Our contribution is the temporal contrast: a larger urban population was associated with higher coverage in both periods, whereas maternal mortality mattered only early and household size and health expenditure only later, and that these characteristics remained relevant after supply constraints eased suggests they reflect durable health-system features rather than transient scarcity. Our models capture these structural correlates but not demand-side barriers such as vaccine hesitancy, which can depress uptake independently of supply^22–28^ and likely became relatively more important as supply eased.

Our findings also speak to how preparedness is measured. Frameworks focused on procurement capture only half the problem: a country can receive doses on time yet remain slow to protect its population without the cold chain, workforce, and community trust needed for administration ^12,29^. A procurement-focused mechanism cannot on its own close coverage gaps rooted in delivery capacity, so speed to coverage, not only doses shipped, belongs in preparedness assessments, a view aligned with a growing literature arguing that preparedness must extend beyond procurement to health-system readiness, sustained financing, and regional coordination^30–35^.

Our findings describe cross-national patterns among countries and territories with reported coverage data, and the structural models describe the smaller complete-case samples of 124 and 157 countries. They should not be extrapolated to subnational populations, to countries without reported coverage, or to routine immunisation programmes outside an emergency rollout.

### Limitations

This study has several limitations. First, our analysis measures inequality rather than inequity. We did not adjust for differences in need across countries, in particular the age structure of the population, which dominates COVID-19 mortality risk. Extending this work to a formal equity analysis that incorporates age-specific risk is an important direction for future research.

Second, COVAX deliveries were not randomly allocated. Allocation reflected country need, diplomatic relationships, and signals of absorptive capacity, so countries receiving earlier or larger deliveries may differ systematically from others in ways that country and time fixed effects cannot fully capture. This limits any causal interpretation of the event-study estimates, which are best read as within-group associations around the timing of delivery rather than as the effect of a randomised intervention. In addition, conventional two-way fixed-effects event-study estimates can be sensitive to heterogeneity in associations across cohorts and event times, which we do not model explicitly.

Third, the event study has limited precision. The standard errors on the post-delivery coefficients range from about 1.2 to 3.4 percentage points, so associations of a few percentage points cannot be excluded. Our result is an absence of evidence for a rapid increase in coverage after the first delivery rather than evidence that no association exists.

Fourth, all analyses were conducted at the country level. Associations between country-level characteristics such as household size or maternal mortality and coverage cannot be interpreted as individual-level relationships, and are subject to the ecological fallacy.

Fifth, reported coverage may be less reliable in lower-capacity settings ^36^. If under-reporting is systematically worse in these countries, the coverage gaps we observe may be conservative rather than exaggerated, since true coverage in lower-capacity settings could be even lower than recorded. Relatedly, the end-of-period comparison of cases and deaths used 2023 data where available and 2022 data otherwise. Because 2023 data were available for a higher share of AMC countries (90.6%) than non-AMC countries (75.4%), this asymmetry may lead to some underestimation of the burden in non-AMC countries. Sixth, national averages can mask substantial subnational inequality by geography, wealth, and gender. Coverage may therefore be overstated for some populations even in countries that nominally reached the 50 percent threshold.

Seventh, the structural models use complete-case data, so the 124 and 157 countries they describe are not a random subset of the 218 in the survival analysis, and countries with incomplete records are likely to differ systematically from those retained.

Finally, the subgroup analyses by health security capacity and WHO region are presented descriptively. The numbers of countries in the highest and lowest capacity groups are small, so we do not attach formal statistical inference to comparisons between these groups.

## Conclusion

Across 218 countries and territories, the speed of the COVID-19 vaccine rollout was strongly unequal, and AMC-supported countries reached 50 percent coverage far more slowly than others. Yet the timing of COVAX deliveries was not, on its own, associated with faster coverage; differences were more consistently associated with country-level structural and health-system characteristics. Delivering vaccines to a country is not the same as protecting its population. Future preparedness efforts should treat speed to coverage as a core measure of vaccine equality and invest in last-mile delivery capacity, trained workforces, and community trust so that supply can be converted into protection when the next emergency arrives.

## Contributors

All authors contributed to the planning, conduct and reporting of the work described in this article. HWL (Hsuan-Wei Lee) conceived and designed the study, developed the analytical framework for the generalized additive mixed model, survival and event study analyses, planned and supervised the conduct of the project, drafted the Introduction and Discussion, and critically revised the manuscript for important intellectual content. YHH (Yi-Hsuan Huang) contributed to the design of the study, acquired and curated the country-level vaccine delivery, sociodemographic and health-system data from public sources, conducted the statistical analyses, produced the tables and figures, and drafted the Methods and Results. TM (Thomas McAndrew) contributed to the conception and design of the study, advised on the epidemiological and statistical interpretation of the findings, and critically revised the manuscript for important intellectual content. All authors had full access to the data, contributed to the interpretation of the results, approved the final version submitted for publication, and agree to be accountable for all aspects of the work. HWL is the guarantor, accepts full responsibility for the finished work and the conduct of the study, had access to the data, and controlled the decision to publish.

Use of artificial intelligence. During the preparation of this revision the authors used generative artificial intelligence tools (Anthropic Claude) to assist with language editing, to check the internal consistency of the reported numbers, cross-references and reporting statements. The tools were not used to collect or generate data, to conduct or interpret the statistical analyses, or to produce any figure or table of results. All analyses were carried out by the authors in R and Python, and all AI-assisted text was checked and revised by the authors, who take full responsibility for the content of the manuscript.

## Funding

This work was supported by The College of Health at Lehigh University (grant number: N/A). The funder had no role in the design of the study, in the collection, analysis or interpretation of the data, in the writing of the report, or in the decision to submit for publication.

## Competing interests

The authors declare no competing financial interests. Author HWL is a co-author of a prior study on COVID-19 vaccine hesitancy in Taiwan. That study is not part of the present analysis and does not affect its conclusions.

## Ethics approval

Ethics approval was not required for this study. All analyses used publicly available, aggregated, country-level secondary data, and no individual participants were involved. Patient consent for publication is not applicable.

## Data availability statement

All data used in this study are publicly available. Country-level vaccination trajectories, cases and deaths were obtained from Our World in Data (https://github.com/owid/covid-19-data); vaccine doses delivered from the UNICEF COVID-19 Vaccine Market Dashboard (https://www.unicef.org/supply/covid-19-market-dashboard); government response indicators from the Oxford COVID-19 Government Response Tracker (https://github.com/OxCGRT/covid-policy-tracker); health expenditure, maternal mortality and education spending from the World Bank World Development Indicators (https://databank.worldbank.org/source/world-development-indicators); preparedness scores from the Global Health Security Index (https://www.ghsindex.org); and synthetic household characteristics from the GLOPOP-S dataset. All sources were accessed between December 2024 and March 2025. The replication code, the derived country-level analysis datasets and accompanying documentation are archived at Zenodo (https://doi.org/10.5281/zenodo.21796606). Redistribution of the original third-party source datasets is restricted by their respective providers, so those files are not included in the archive; instructions for retrieving them are given in the repository README.

## Notes

### Competing Interest Statement

The authors have declared no competing interest.

