## Supplementary Material for "Temporal inequalities in the global COVID-19 vaccine rollout: a cross-national observational study of delivery, health-system capacity, and time to coverage"

### Supplementary File

This file contains supplementary figures, tables, model specifications and the STROBE checklist for the manuscript "Temporal inequalities in the global COVID-19 vaccine rollout".

#### Supplementary Figures

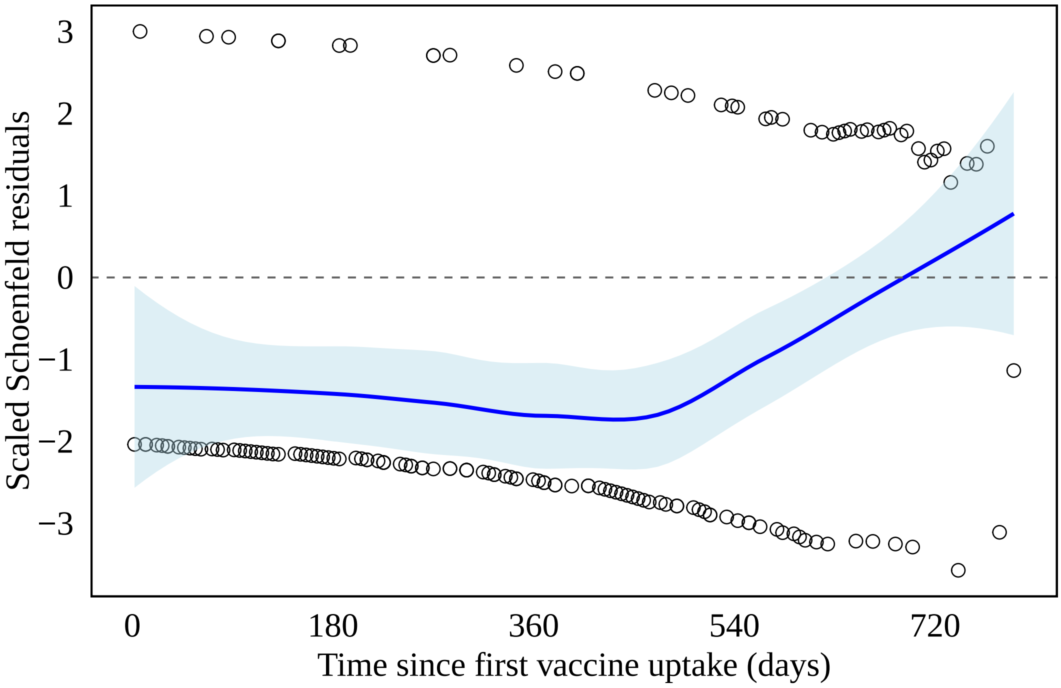


Figure S1: Schoenfeld residual plot assessing the proportional hazards assumption for AMC status. The scaled Schoenfeld residuals (open circles) are expected to fluctuate randomly around zero under the assumption of proportional hazards. The solid line represents a smoothed estimate with 95% confidence intervals (shaded area), and the dashed line indicates the $y = 0$ reference line.

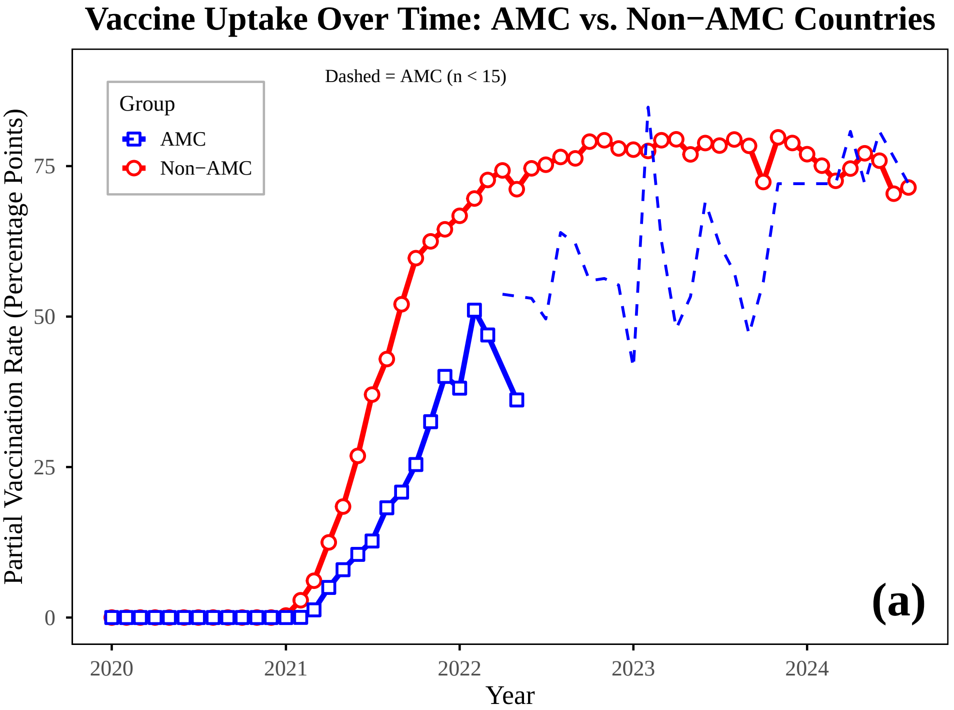


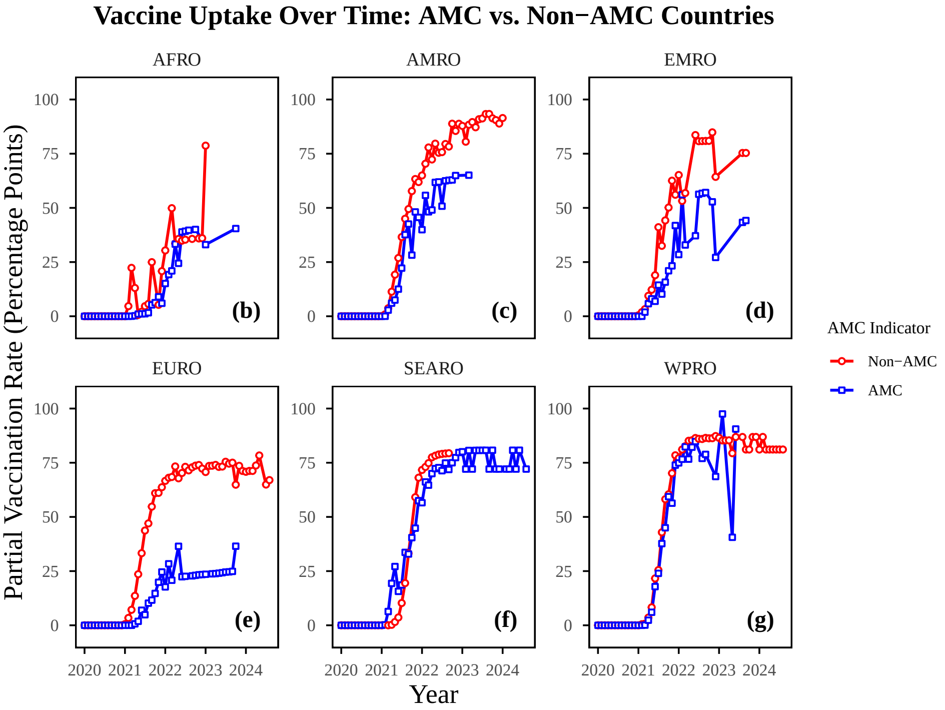


Figure S2: Vaccine uptake over time in AMC vs. Non-AMC countries. (a) shows the vaccination trends for all countries. (b)-(g) further stratifies countries by WHO regions: the African Region (AFRO), the Region of the Americas (AMRO), the Eastern Mediterranean Region (EMRO), the European Region (EURO), the South-East Asia Region (SEARO), and the Western Pacific Region (WPRO). For subgroups with <15 countries at a given time point, dashed lines are used to indicate limited sample size to avoid overinterpretation.

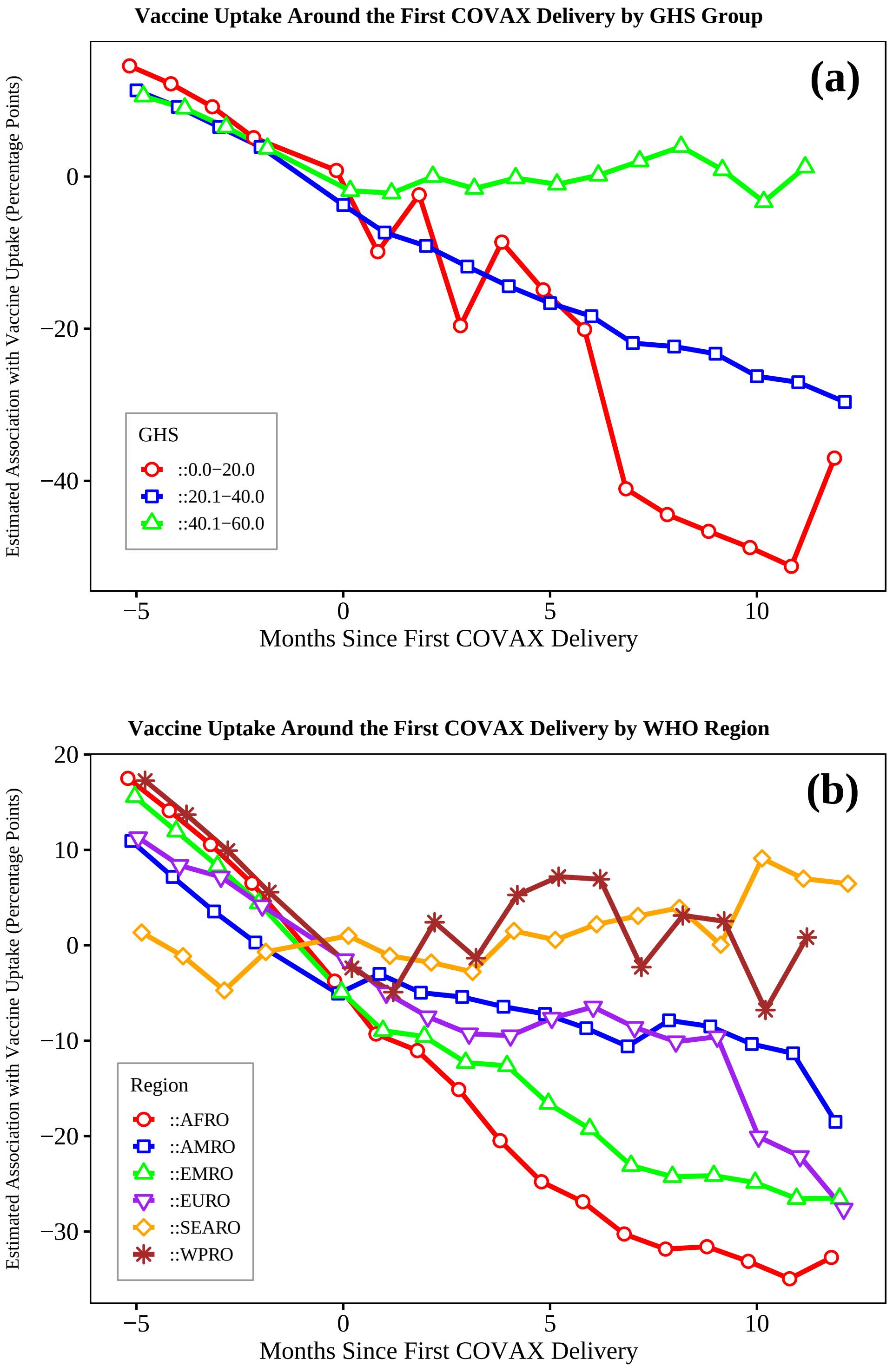


Figure S3: Estimated association between the timing of the first COVAX delivery and national COVID-19 vaccine uptake, by Global Health Security (GHS) Index group and WHO Region. Because the low- and high-GHS groups contain only four and seven countries, respectively, these subgroup trajectories are descriptive and should not be interpreted as formal evidence of between-group heterogeneity. (a) AMC countries are classified into three groups based on their GHS Index scores (0.0–20.0, 20.1–40.0, and 40.1–60.0). (b) AMC countries are grouped by WHO Region, including the African Region (AFRO), Region of the Americas (AMRO), Eastern Mediterranean Region (EMRO), European Region (EURO), South-East Asia Region (SEARO), and Western Pacific Region (WPRO). All values are reported in percentage points of coverage. Unlike Figure 3 in the main text, the subgroup trajectories are not centred on the reference month, so their values at a given event time are not directly comparable with the coefficients in Figure 3 in the main text and should be read only as within-subgroup descriptive patterns.

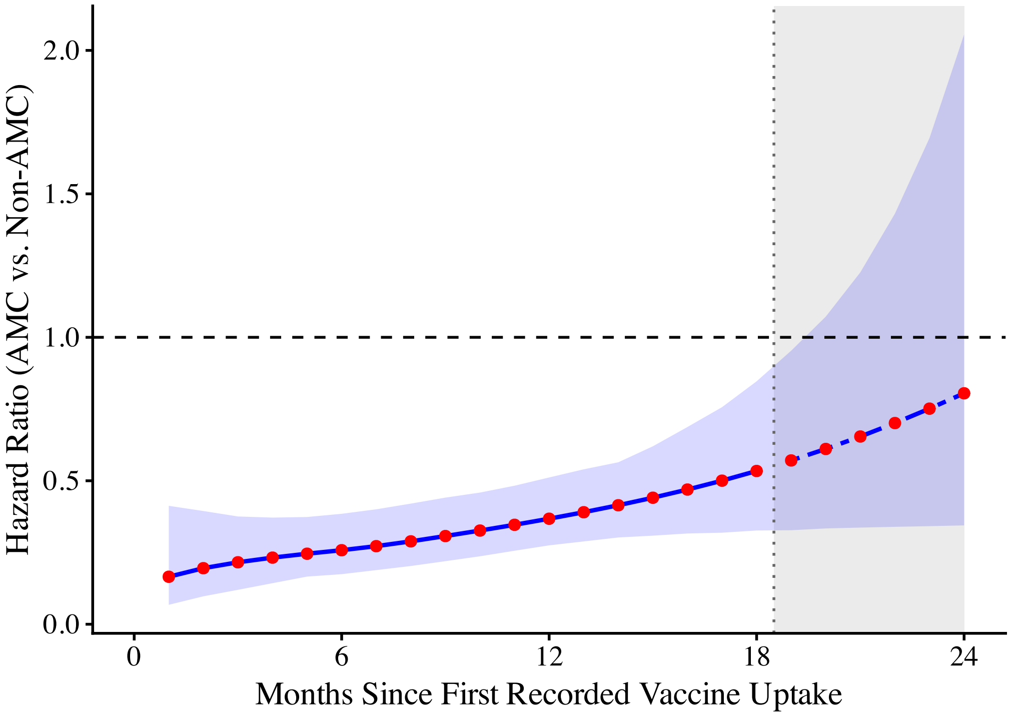


Figure S4: Estimated time-varying hazard ratio of reaching 50% vaccine uptake for AMC versus non-AMC countries. The hazard ratio was estimated from the Royston-Parmar flexible parametric survival model with a time-varying coefficient for AMC status. Values below 1 indicate a lower hazard of reaching the vaccination threshold among AMC countries compared with non-AMC countries, i.e., slower attainment rather than lower effectiveness. The gradual increase in $HR(t)$ over time indicates attenuation of the initial difference in vaccine rollout timing. Shaded bands denote 95% confidence intervals derived from 1,000 bootstrap replicates of the model parameters. Estimates beyond month 18 (shaded grey) are shown for reference only, as the non-AMC group's risk set became small with few accrued events after this point, resulting in unstable hazard estimates. Time is measured from the start of follow-up, that is from a country's first recorded vaccine uptake, and the horizontal axis in the plot refers to this follow-up time.

### Supplementary Tables 6

Table S1: Proportional hazards assumption test using Schoenfeld residuals

| **Variable** | **Chi-squared** | **df** | **p-value** |
|---|---|---|---|
| AMC_IND | 6.40 | 1 | 0.011 |
| GLOBAL | 6.40 | 1 | 0.011 |

| Group | Count |
|---|---|
| AMC | 88 |
| Non-AMC | 130 |

Table S2: Number of AMC and Non-AMC countries

Table S3: Survival Analysis log-rank test

| Group | N | Observed | Expected | $(O-E)^2/E$ | $(O-E)^2/V$ |
|---|---|---|---|---|---|
| Non-AMC | 130 | 108 | 68.9 | 22.2 | **42.9*****|
| AMC | 88 | 45 | 84.1 | 18.2 | **42.9***** |

Note: $O$ denotes observed and $E$ expected events, that is countries reaching 50% coverage, under the null hypothesis of no difference between groups. The final column is the log-rank test statistic, $\chi^2_1 = 42.9$, $p = 6 \times 10^{-11}$. *** $p < 0.001$.

Table S4: Survival Analysis, after 365.25 days

| Group | N Risk | N Event | Surv. Prob. | Std. Error | Lower 95% CI | Upper 95% CI |
|---|---|---|---|---|---|---|
| Non-AMC | 26.0 | 102.0 | 0.211 | 0.036 | 0.151 | 0.295 |
| AMC | 58.0 | 30.0 | 0.659 | 0.051 | 0.567 | 0.766 |

Note: Values are Kaplan–Meier estimates at 365.25 days of follow-up. Surv. Prob. is the estimated proportion of countries that had not yet reached 50% coverage at that time. N Risk and N Event are the numbers still at risk and the numbers that had reached the threshold by that time.

### Supplementary Appendix

### 1. Appendix

#### 1.1 Model Robustness Checks

This appendix presents supporting material for the models at the two stages, July 2021 and the later stage (2023). For each stage, diagnostic plots are provided in Figures S5 and S6, including Normal Q-Q plot, residuals vs. linear predictor plot, histogram of residuals, and response vs. fitted values plot. Tests for heteroscedasticity and residual normality are also reported. Residual normality is not an assumption of a quasi-binomial model, so these tests are provided for completeness and are not treated as tests of model validity.

##### 1.1.1 July 2021 Stage (GAMM)

**Breusch-Pagan Test for Heteroscedasticity:**

```
studentized Breusch-Pagan test
```

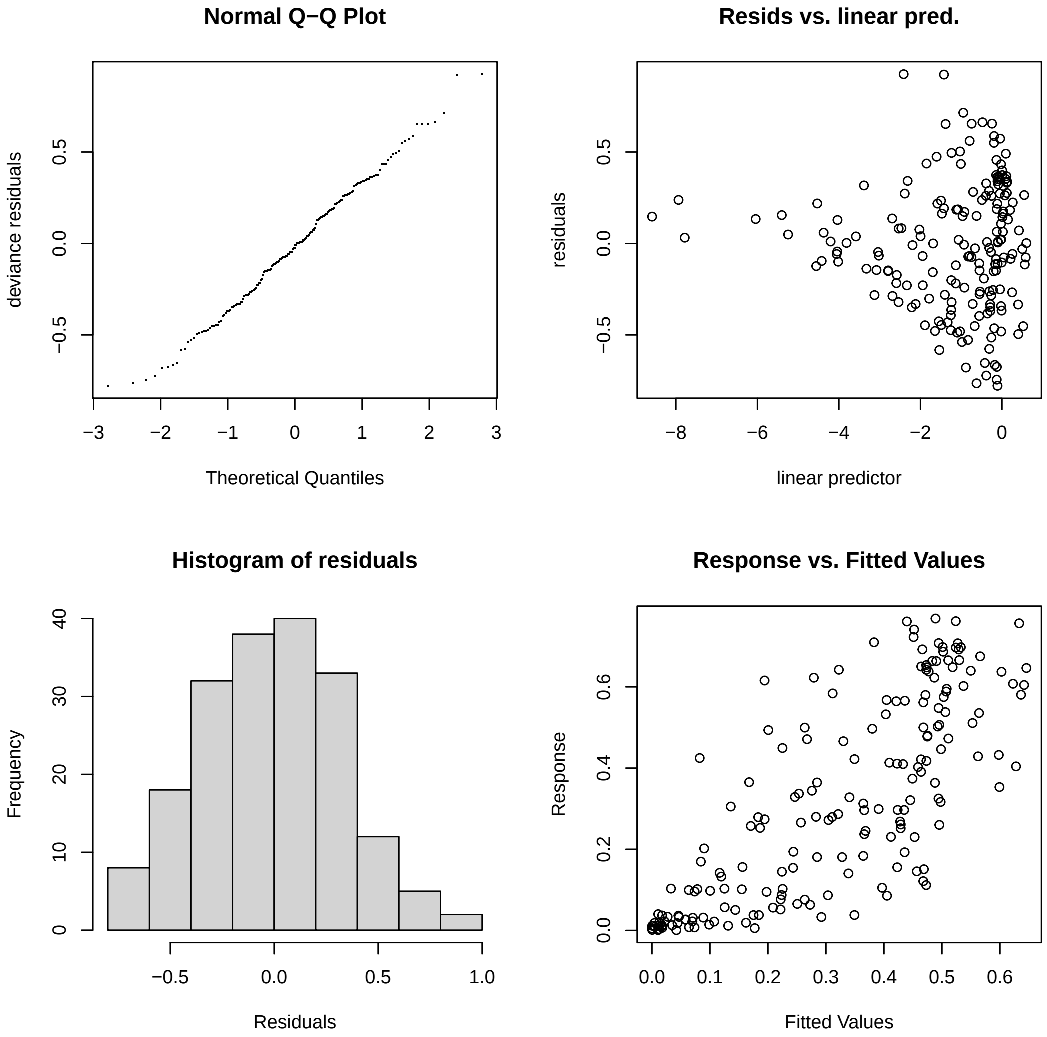


Figure S5: Diagnostic plots for the July 2021 stage model

```
data:  model_partial_distinct
BP = 23.584, df = 14, p-value = 0.05141
```

##### Shapiro-Wilk Test for Normality of Residuals:

```
Shapiro-Wilk normality test
data:  residuals(model_partial_distinct, type = "deviance")
W = 0.98986, p-value = 0.4969
```

##### 1.1.2 Later Stage (2023) GAMM

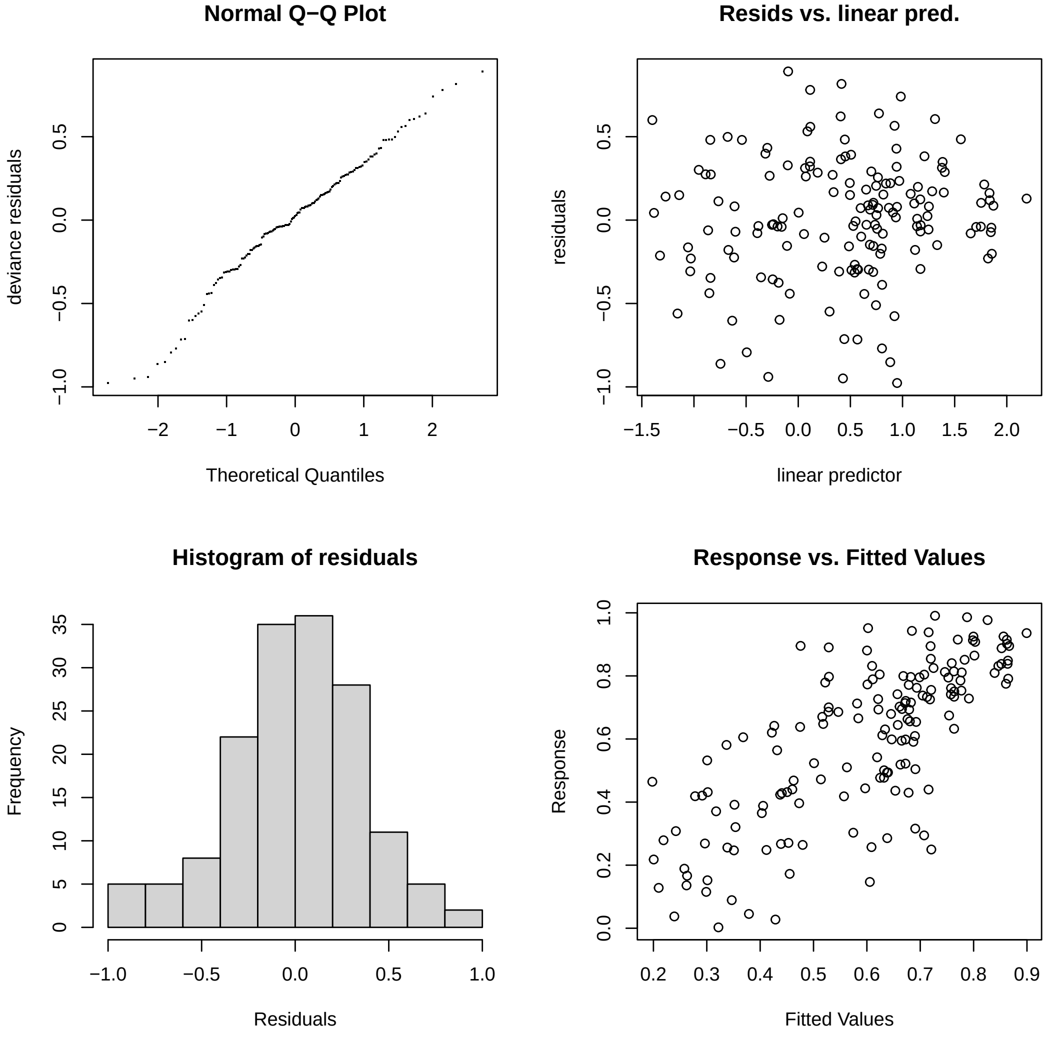


Figure S6: Diagnostic plots for the later stage (2023) model

##### Breusch-Pagan Test for Heteroscedasticity:

```
studentized Breusch-Pagan test
data:  model_partial_distinct
BP = 20.782, df = 18, p-value = 0.2906
```

##### Shapiro-Wilk Test for Normality of Residuals:

```
Shapiro-Wilk normality test
data:  residuals(model_partial_distinct, type = "deviance")
W = 0.98382, p-value = 0.06336
```

##### 1.1.3 Survival Analysis

Table S5: Flexible parametric (Royston-Parmar) survival model with a time-varying coefficient for AMC status

| Variable / Parameter | Coefficient (Log HR) | Std. Error | Hazard Ratio (HR) | 95% CI for HR |
|---|---|---|---|---|
| **Core Covariate & Time-Varying Term** | | | | |
| AMC-supported status ($AMC\_IND$) | -3.075 | 0.959 | 0.046 | [0.007, 0.303] |
| $\gamma_1 \times$ AMC-supported status | 0.300 | 0.147 | 1.350 | [1.011, 1.803] |
| **Baseline Hazard Splines** | | | | |
| $\gamma_0$ (Intercept) | -5.640 | 1.176 | – | – |
| $\gamma_1$ | 0.634 | 0.323 | – | – |
| $\gamma_2$ | -1.129 | 0.162 | – | – |
| $\gamma_3$ | 1.317 | 0.177 | – | – |
| **Model Diagnostics** | | | | |
| Observations ($N$) = 218 \| Number of Events = 153 | | | | |
| Log-Likelihood = -1067.168 \| AIC = 2146.337 | | | | |

*Note:* The model utilizes restricted cubic splines with two internal knots ($k = 2$) on the hazard scale, incorporating a time-varying coefficient for AMC status via interaction with the first spline basis function ($\gamma_1$). Due to the time-varying nature, the net hazard ratio $HR(t)$ changes over time; point estimates and $HR(t)$ trajectories are detailed in Figure S4.

#### 1.2 Model Specifications

##### 1.2.1 Generalized additive mixed models

To identify structural correlates across different periods, Generalized Additive Mixed Models (GAMMs) were estimated for both the early stage (2021) and the later stage of the pandemic using a `quasibinomial` family and a `logit` link function.

##### Model 1: Early Stage Baseline Model (2021)

In the early stage model, the adjusted partial vaccine uptake rate $Y_i$ was utilized directly in its original scale as the dependent variable. Let $\mu_{i,\text{early}} = E(Y_i)$, where $Y_i \in [0, 1]$. The specification is formulated as follows:

$$
\begin{aligned}
\ln\left(\frac{\mu_{i,\text{early}}}{1-\mu_{i,\text{early}}}\right) = {} & \beta_0 + \beta_1 \ln(\text{HHSize}_i + 1) + \beta_2 \ln(\text{Educ}_i + 1) + \beta_3 \ln(\text{Pop}_i + 1) \\
& + \beta_4 \ln\left(\sqrt{\text{PopDens}_i + 1}\right) + \beta_5 \ln(\text{Stringency}_i + 1) + \beta_6 \text{MaternalMort}_i \\
& + \beta_7 \sqrt{\text{GovEdShare}_i} + \beta_8 \ln(\text{UrbanPop}_i + 1) + \beta_9 \text{PC1}_i \\
& + s(\text{WHORegion}_i, \text{bs} = \text{'re'}) \qquad (1)
\end{aligned}
$$

##### Model 2: Later Stage Interaction Model

In the later stage model the dependent variable was again the adjusted partial vaccination share on its original scale. Let $\mu_{i,\text{later}} = E(Y_i)$, where $Y_i \in [0,1]$. The specification incorporating context-specific interaction effects is formulated as follows:

$$
\begin{aligned}
\ln\left(\frac{\mu_{i,\text{later}}}{1-\mu_{i,\text{later}}}\right) = {} & \gamma_0 + \gamma_1 \text{HHType}_i + \gamma_2 \ln(\text{HHSize}_i + 1) + \gamma_3 \ln(\text{Educ}_i + 1) \\
& + \gamma_4 \ln(\text{UrbanPop}_i + 1) + \gamma_5 \ln\left(\sqrt{\text{PopDens}_i + 1}\right) + \gamma_6 \ln(\text{HealthExp}_i + 1) \\
& + \gamma_7 \left[\ln\left(\sqrt{\text{PopDens}_i + 1}\right) \times \ln(\text{HealthExp}_i + 1)\right] \\
& + \gamma_8 \ln(\text{Stringency}_i + 1) + \gamma_9 \left[\ln\left(\sqrt{\text{PopDens}_i + 1}\right) \times \ln(\text{Stringency}_i + 1)\right] \\
& + \gamma_{10} \sqrt{\text{GovEdShare}_i} + \gamma_{11} \ln(\text{RuralMean}_i + 1) \\
& + \gamma_{12} \left[\sqrt{\text{GovEdShare}_i} \times \ln(\text{RuralMean}_i + 1)\right] \\
& + s(\text{HHType}_i, \text{WHORegion}_i, \text{bs} = \text{'re'}) \qquad (2)
\end{aligned}
$$

Where the model parameters and variables are defined below:

- $\beta_0, \gamma_0$: Structural intercepts for the early and later stage models, respectively.
- $\text{HHType}_i$: Household type categorical indicator consisting of categories 3, 6, and 7. In Model 2, it is evaluated as a set of dummy variables where the standard "couple with children" category (`HHTYPE_MODE3`) serves as the omitted reference group. Individual parametric coefficients are estimated for "couple with children and (non-)relatives" (`HHTYPE_MODE6`) and "one parent with children and (non-)relatives" (`HHTYPE_MODE7`).
- $\ln(\text{HHSize}_i + 1)$: Dominant household size category (log).
- $\ln(\text{Educ}_i + 1)$: Dominant educational attainment level (log).
- $\ln(\text{Pop}_i + 1)$: Total population (log).

- $V1_i$: Vaccine prioritisation ranking (VPR), the rank order of population groups eligible for vaccination. This variable does not enter Model 1 or Model 2. It appears only in the sensitivity specification reported in Table S7, which adds $\beta V1_i$ to Equation 1 and changes nothing else. It was excluded from the main models because a country typically reaches universal eligibility only after vaccinating its priority groups, so the variable is potentially endogenous to uptake.
- $PC1_i$: Principal component representing COVAX participation and supply characteristics.
- $\ln(\text{UrbanPop}_i + 1)$: Urban population (log).
- $\ln\left(\sqrt{\text{PopDens}_i + 1}\right)$: Population density (log).
- $\ln(\text{Stringency}_i + 1)$: Government response stringency (log).
- $\text{MaternalMort}_i$: Maternal mortality ratio.
- $\ln(\text{HealthExp}_i + 1)$: Health expenditure (log).
- $\sqrt{\text{GovEdShare}_i}$: Public spending on education (% of GDP, sqrt).
- $\ln(\text{RuralMean}_i + 1)$: Rural population mean (log).
- $\ln\left(\sqrt{\text{PopDens}_i + 1}\right) \times \ln(\text{HealthExp}_i + 1)$: Interaction term between population density and health expenditure.
- $\ln\left(\sqrt{\text{PopDens}_i + 1}\right) \times \ln(\text{Stringency}_i + 1)$: Interaction term between population density and government stringency.
- $\sqrt{\text{GovEdShare}_i} \times \ln(\text{RuralMean}_i + 1)$: Interaction term between public spending on education and rural population mean.
- $s(\cdot)$: Random effect smooth terms accounting for geographic-level heterogeneity; the later-stage model additionally includes crossed random effects between household type and WHO region.

##### 1.2.2 Event study

$$Y_{it} = \alpha_i + \lambda_t + \sum_{k=-m}^{q} \beta_k D_{it}^k + \epsilon_{it} \tag{3}$$

Where $Y_{it}$ is the partial-vaccination coverage of country $i$ in month $t$, in percentage points, taken as the last value recorded in that month. $\alpha_i$ and $\lambda_t$ represent country and calendar month-year fixed effects, respectively, to control for unobserved time-invariant characteristics and global temporal shocks. $D_{it}^k$ denotes the event time dummy variables, indicating $k$ months relative to the first COVAX delivery in country $i$. The indices $-m$

and $q$ define the bounds of the temporal observation window for capturing pre-treatment trends and post-treatment dynamic associations, respectively. $\epsilon_{it}$ is the error term.

For the event study analysis, a two-way fixed effects model was employed to estimate the dynamic associations between COVAX deliveries and national vaccine uptake. The model specification included event time dummy variables , country fixed effects to control for time-invariant unobserved heterogeneity , and calendar month-year fixed effects to account for global temporal trends. To fully capture potential anticipatory trends and long-term dynamic deployment patterns, the temporal window was restricted to 5 months prior to and 12 months following the initial delivery (i.e., setting $m = 5$ and $q = 12$ in the summation). The month immediately preceding the delivery ($k = -1$) was omitted from the estimation and designated as the reference period to avoid perfect multicollinearity. No additional time-varying covariates were included in this model.

##### 1.2.3 Survival analysis

$$h(t|\mathrm{AMC}_i) = h_0(t)\exp(\beta \cdot \mathrm{AMC}_i) \tag{4}$$

Where $h_0(t)$ represents the baseline hazard function, and $\mathrm{AMC}_i$ is a binary indicator denoting the country's Advance Market Commitment status (1 if AMC-supported, 0 if non-AMC). $\beta$ is the coefficient for AMC status on the log hazard scale.

A univariate Cox proportional hazards model with AMC status as the sole explanatory variable was fitted to test the proportional hazards assumption using Schoenfeld residuals. Because that assumption was violated, we do not report a single proportional hazards ratio; the hazard ratios reported in the main text come from the flexible parametric model with a time-varying AMC coefficient, and the group comparison is summarised by the log-rank test and the Kaplan–Meier estimates.

Table S6: Variable Classification, Theoretical Justifications, and Collinearity Diagnostics

| **Dimension** | **Theoretical Justification & Variables Included** | **Collinearity Status** |
|---|---|---|
| **1. Socio-Demographic** | Controls for demand-side frictions and viral exposure risks based on household living structures. Includes *household size category* (crowding proxy) and complex family structures (*extended or single-parent households*). | Stable (Max VIF = 4.26) |
| **2. Healthcare Capacity** | Proxies infrastructure readiness and institutional capacity to execute mass campaigns. Includes *maternal mortality ratio* and *health expenditure* (system quality), along with *stringency index.* | Stable (Max VIF = 3.15) |
| **3. Human Capital** | Captures health literacy and demand-side acceptance. Higher *educational attainment levels* and *education spending* proxy a population's capacity to counter vaccine hesitancy and misinformation. | Stable (Max VIF = 3.61) |
| **4. Spatial Geography** | Controls for scale and "last-mile" logistical barriers. Urban components (*urban population*, *density*) proxy centralized distribution ease, while *rural share* captures remote delivery frictions. | Stable (Max VIF = 2.26) |
| **5. Global Supply** | Captures external resource constraints. Highly correlated supply vectors (*AMC eligibility*, *post-AMC timeline*, *lagged cumulative deliveries*) were synthesized into a single composite metric (**PC1**) via PCA. | Structurally Eliminated (PC1 Variance = 72.1%) |

*Note:* Reported VIF values are the Stage 2 baseline estimates, which are the larger of the two stages; the corresponding Stage 1 maximum was 3.82. Reported VIF values represent baseline estimates excluding multiplicative product terms. Higher VIF values for the multiplicative product terms are expected in models that include interactions and were not, by themselves, interpreted as evidence of problematic collinearity; main-effect coefficients are interpreted conditionally on the corresponding interaction terms.

Table S7: Sensitivity Analysis: GAMM Results for Partial COVID-19 Vaccine Uptake Including the Vaccine Prioritization Ranking (VPR) Variable (July 2021). This specification is presented for comparison only; the VPR variable was excluded from the main model reported in the text because it is potentially endogenous to uptake.

| **Variable** | **Estimate** | **Std. Error** | **t value** | **p-value** | **AME [95% CI]** |
|---|---|---|---|---|---|
| (Intercept) | -2.306 | 2.872 | -0.803 | 0.424 | — |
| Dominant household size category (log, +1) | -1.807 | 1.146 | -1.577 | 0.118 | -0.288<br>[-0.606, 0.031] |
| Dominant educational attainment level (log, +1) | 0.460 | 0.317 | 1.454 | 0.149 | 0.073<br>[-0.033, 0.179] |
| Total population (log, +1) | -0.063 | 0.049 | -1.294 | 0.198 | -0.010<br>[-0.025, 0.005] |
| Population density (log, sqrt, +1) | 0.153 | 0.117 | 1.311 | 0.192 | 0.024<br>[-0.012, 0.061] |
| Government response stringency (log, +1) | 0.172 | 0.242 | 0.714 | 0.477 | 0.027<br>[-0.048, 0.103] |
| Maternal mortality ratio | -0.005 | 0.002 | -3.127 | **0.002**** | **-0.0008****<br>**[-0.0013, -0.0002]** |
| Public spending on education (% of GDP, sqrt) | 0.075 | 0.222 | 0.336 | 0.737 | 0.012<br>[-0.057, 0.081] |
| Urban population (log, +1) | 0.667 | 0.305 | 2.189 | **0.031*** | **0.106***<br>**[0.010, 0.203]** |
| Vaccine prioritization ranking (VPR) | 0.395 | 0.154 | 2.563 | **0.012*** | **0.063****<br>**[0.016, 0.110]** |
| COVAX participation and supply (PC1) | 0.007 | 0.067 | 0.111 | 0.912 | 0.001<br>[-0.020, 0.022] |
| *Smooth term (Random effect)* | | | | | |
| WHO regional | | edf = 0.675 | F = 0.153 | 0.335 | — |

Adjusted $R^2 = 0.608$, Deviance explained = 64.5%, Scale est. = 0.12364, $n = 124$.
Note: The Estimate, Std. Error, t value and p-value columns refer to the parametric coefficients on the logit scale. Average marginal effects (AME) give the average change in the predicted probability of partial vaccine uptake (0–1 scale) for a one-unit increase in the covariate as it enters the model, that is in its log-transformed or square-root-transformed value. AME standard errors and 95% confidence intervals [CI] are obtained by the delta method, and the interval rather than a separate p-value summarises AME precision. Significance markers: . $p < 0.10$, * $p < 0.05$, ** $p < 0.01$, *** $p < 0.001$.

### STROBE Statement—Checklist of items that should

Table S8: Comparison of Event Study Regressions

| | (a) Clean Monthly (Last) | (b) Daily + Date FE | (c) Daily + Month FE |
|---|---|---|---|
| Dependent Var.: | PARTIALLY_VACCINATED_ADJ | | |
| **Pre-treatment Periods** | | | |
| Month $t = -5$ | 3.986 (6.117) | 3.871 (6.126) | −6.263 (4.934) |
| Month $t = -4$ | 3.268 (4.554) | 2.926 (5.271) | −4.057 (3.942) |
| Month $t = -3$ | 2.398 (2.873) | 2.214 (3.161) | −0.959 (2.537) |
| Month $t = -2$ | 1.720 (1.393) | 1.369 (2.326) | 1.616 (2.111) |
| Month $t = -1$ | | *Omitted Reference Category* | |
| **Post-treatment Periods** | | | |
| Month $t = 0$ | −1.237 (1.210) | −0.876 (2.111) | 0.886 (1.944) |
| Month $t = 1$ | −2.854 (1.814) | −2.679 (2.578) | 2.839 (2.318) |
| Month $t = 2$ | −1.671 (2.353) | −4.150 (3.366) | 4.112 (2.826) |
| Month $t = 3$ | −2.827 (2.743) | −4.725 (3.774) | $6.252^{*}$ (3.048) |
| Month $t = 4$ | −2.120 (2.963) | −6.299 (4.008) | $7.240^{*}$ (3.256) |
| Month $t = 5$ | −2.071 (3.008) | $-6.748^{\cdot}$ (3.917) | $9.559^{**}$ (3.319) |
| Month $t = 6$ | −1.529 (2.971) | −6.250 (3.903) | $12.622^{***}$ (3.433) |
| Month $t = 7$ | −3.263 (3.395) | −5.606 (4.031) | $15.503^{***}$ (3.458) |
| Month $t = 8$ | −1.431 (3.007) | −5.975 (4.293) | $17.702^{***}$ (3.448) |
| Month $t = 9$ | −0.433 (2.714) | −5.967 (4.910) | $20.303^{***}$ (3.684) |
| Month $t = 10$ | −1.676 (2.473) | −8.179 (5.325) | $20.863^{***}$ (3.370) |
| Month $t = 11$ | −0.485 (1.917) | $-10.501^{\cdot}$ (5.838) | $21.492^{***}$ (3.378) |
| Month $t = 12$ | *Dropped* | −9.299 (6.365) | $24.793^{***}$ (3.424) |
| **Fixed Effects** | | | |
| ENTITY | Yes | Yes | Yes |
| YEARMONTH | Yes | No | Yes |
| VACCINED_DATE | No | Yes | No |
| Observations | 1,344 | 9,644 | 9,644 |
| $R^2$ | 0.71584 | 0.84668 | 0.83477 |
| Within $R^2$ | 0.00552 | 0.00687 | 0.01559 |

*Notes:* Standard errors in parentheses are clustered at the country (`ENTITY`) level. In Column (a), Month 12 is dropped automatically due to exact collinearity with the year-month fixed effects. Column (a) is the main specification and Column (b) is a daily-frequency robustness check. Column (c) reproduces the misspecified model from the original submission, in which daily observations were combined with monthly fixed effects. It is shown only to document how that combination generated artificially precise and significant post-delivery estimates, and it should not be interpreted as a valid robustness specification.

*Sample Power Distribution:* The number of unique countries contributing to each event-time coefficient is 75 for months −5 to 6 and 74 for months 7 onward. In Column (a) month 12 is dropped because of exact collinearity with the year-month fixed effects, so 74 countries contribute to months 7 to 11; in Columns (b) and (c), 74 countries also contribute at month 12.

#### be included in reports of cross-sectional studies

Page numbers are generated automatically from the compiled manuscript, and section names are given alongside them so that each item remains locatable if the pagination changes in production.

| Item No | Recommendation | Location in Manuscript |
|---|---|---|
| **Title and abstract** | | |
| 1 (a) | Indicate the study's design with a commonly used term in the title or the abstract | Title; Abstract, Methods (p. 1) |
| 1 (b) | Provide in the abstract an informative and balanced summary of what was done and what was found | Abstract (p. 1) |
| **Introduction** | | |
| 2 | **Background/rationale:** Explain the scientific background and rationale for the investigation being reported | Introduction |
| 3 | **Objectives:** State specific objectives, including any prespecified hypotheses | Introduction, final paragraph |
| **Methods** | | |
| 4 | **Study design:** Present key elements of study design early in the paper | Abstract, Methods (p. 1); Methods, opening paragraphs |
| 5 | **Setting:** Describe the setting, locations, and relevant dates, including periods of recruitment, exposure, follow-up, and data collection | Methods 2.4, Data Sources and Collection; Methods 2.2, follow-up definition |
| 6 (a) | **Participants:** Give the eligibility criteria, and the sources and methods of selection of participants | Methods 2.2, AMC classification; Methods 2.3, event-study sample; Methods 2.4, exclusions; Figure 1 in the main text |
| 7 | **Variables:** Clearly define all outcomes, exposures, predictors, potential confounders, and effect modifiers. Give diagnostic criteria, if applicable | Methods 2.1, 2.2, 2.3 and 2.4; Appendix, Model Specifications |
| 8 | **Data sources/measurement:** For each variable of interest, give sources of data and details of methods of assessment (measurement). Describe comparability of assessment methods if there is more than one group | Methods 2.4, Data Sources and Collection |

*Continued on next page*

– *Continued from previous page*

| Item No | Recommendation | Location in Manuscript |
| --- | --- | --- |
| 9 | **Bias:** Describe any efforts to address potential sources of bias | Methods 2.1, principal component analysis and variance inflation factors; Methods 2.2, proportional hazards assessment; Methods 2.3, pre-treatment coefficients; Methods 2.4, missing data; Discussion, Limitations |
| 10 | **Study size:** Explain how the study size was arrived at | Methods 2.4; Figure 1 in the main text |
| 11 | **Quantitative variables:** Explain how quantitative variables were handled in the analyses. If applicable, describe which groupings were chosen and why | Methods 2.1, transformations and composite indicator; Methods 2.3, GHS bands; Appendix, Model Specifications |
| 12 (a) | **Statistical methods:** Describe all statistical methods, including those used to control for confounding | Methods 2.1–2.3; Appendix, Model Specifications |
| 12 (b) | Describe any methods used to examine subgroups and interactions | Methods 2.3, WHO region and GHS strata; Results 3.1, interaction terms; Appendix, Equation 2 |
| 12 (c) | Explain how missing data were addressed | Methods 2.4, complete-case criteria |
| 12 (d) | If applicable, describe analytical methods taking account of sampling strategy | Not applicable. Aggregated country-level secondary data analysed on a complete-case basis (Methods 2.4, Data Sources and Collection) |
| 12 (e) | Describe any sensitivity analyses | Methods 2.4, vaccine prioritisation variable; Appendix, Table S7 and Table S8 |
| **Results** | | |
| 13 (a) | **Participants:** Report numbers of individuals at each stage of study eg numbers potentially eligible, examined for eligibility, confirmed eligible, included in the study, completing follow-up, and analysed | Methods 2.4; Figure 1 in the main text; Results 3.3 |
| 13 (b) | Give reasons for non-participation at each stage | Methods 2.4, exclusions by reason; Figure 1 in the main text |

*Continued on next page*

– *Continued from previous page*

| Item No | Recommendation | Location in Manuscript |
|---|---|---|
| 13 (c) | Consider use of a flow diagram | Figure 1 in the main text, analytic sample for each component of the study |
| 14 (a) | **Descriptive data:** Give characteristics of study participants (eg demographic, clinical, social) and information on exposures and potential confounders | Methods 2.4, variables and sources; Table S2; Table 1 in the main text; Appendix, Table S6. The distribution of each individual covariate is not tabulated |
| 14 (b) | Indicate number of participants with missing data for each variable of interest | Methods 2.4, complete-case criteria and resulting sample sizes; Figure 1 in the main text. Counts are reported per analysis rather than per variable |
| 15 | **Outcome data:** Report numbers of outcome events or summary measures | Table 1 in the main text; Results 3.3, events and censoring; Table S3; Table S4; Table 4 in the main text |
| 16 (a) | **Main results:** Give unadjusted estimates and, if applicable, confounder-adjusted estimates and their precision (eg, 95% confidence interval). Make clear which confounders were adjusted for and why they were included | Table 2 in the main text; Table 3 in the main text; Table 4 in the main text; Tables S3 and S4; Figure S4; Figure 2 in the main text |
| 16 (b) | Report category boundaries when continuous variables were categorized | Methods 2.3, Global Health Security Index bands of 0–20, 20.1–40 and 40.1–60 |
| 16 (c) | If relevant, consider translating estimates of relative risk into absolute risk for a meaningful time period | Results 3.3. The time-varying hazard ratios are reported alongside the absolute Kaplan–Meier probabilities of not having reached 50% coverage at one year, in Table S4 |

*Continued on next page*

– *Continued from previous page*

| Item No | Recommendation | Location in Manuscript |
|---|---|---|
| 17 | **Other analyses:** Report other analyses done eg analyses of subgroups and interactions, and sensitivity analyses | Results 3.2, subgroup trajectories; Results 3.3, flexible parametric model and Wilcoxon rank-sum tests; Appendix, Table S7 and Table S8 |
| **Discussion** | | |
| 18 | **Key results:** Summarise key results with reference to study objectives | Discussion, opening paragraphs |
| 19 | **Limitations:** Discuss limitations of the study, taking into account sources of potential bias or imprecision. Discuss both direction and magnitude of any potential bias | Discussion: Limitations |
| 20 | **Interpretation:** Give a cautious overall interpretation of results considering objectives, limitations, multiplicity of analyses, results from similar studies, and other relevant evidence | Discussion; Conclusion |
| 21 | **Generalisability:** Discuss the generalisability (external validity) of the study results | Discussion, final paragraph; Limitations |
| **Other information** | | |
| 22 | **Funding:** Give the source of funding and the role of the funders for the present study and, if applicable, for the original study on which the present article is based | Funding statement |
